# The COVID-19 Yorkshire Rehabilitation Scale (C19-YRS): application and psychometric analysis in a post-COVID-19 syndrome cohort

**DOI:** 10.1101/2021.06.28.21259613

**Authors:** Nick Preston, Amy Parkin, Sophie Makower, Denise H Ross, Jeremy Gee, Stephen J Halpin, Mike C Horton, Rory J O’Connor, Manoj Sivan

## Abstract

**Background:** As our understanding of the nature and prevalence of Post-COVID-19 syndrome (PCS) is increasing, a measure of the impact of COVID-19 could provide valuable insights into patients’ perceptions in clinical trials and epidemiological studies, as well as routine clinical practice.

**Objective:** To evaluate the clinical usefulness and psychometric properties of the COVID-19 Yorkshire Rehabilitation Scale (C19-YRS) in patients with PCS.

**Design:** A prospective, observational study of 187 consecutive patients attending a post-COVID-19 rehabilitation clinic. The C19-YRS was used to record patients’ symptoms, functioning and disability. A global health question was used to measure the overall impact of PCS on health. Classical psychometric methods (data quality, scaling assumptions, targeting, reliability and validity) were used to assess the C19-YRS.

**Results:** For the overall scale, missing data were low, scaling and targeting assumptions were satisfied, and internal consistency was high (Cronbach’s alpha = 0.891). Relationships between perception of health and patients’ reports of symptoms, functioning and disability demonstrated good concordance.

**Conclusions:** This is the first study to examine the psychometric properties of an outcome measure in patients with PCS. In this sample of patients, the C19-YRS was clinically useful and satisfied standard psychometric criteria, providing preliminary evidence of its suitability as a measure of PCS.

## Introduction

The medium and long-term problems experienced by survivors of COVID-19 are emerging, but standardized assessments of functioning, disability and health are lacking. Long-term symptoms of COVID-19 might be predicted from the previous coronavirus outbreaks in 2002 and 2012 – Severe Acute Respiratory Syndrome (SARS) and Middle East Respiratory Syndrome (MERS) respectively. A meta-analysis of follow-up studies demonstrated that 25% of hospitalized survivors of SARS and MERS experienced reduced lung function and lower exercise capacity six months post-discharge.^1^ One year on, post-traumatic stress disorder (PTSD), depression, anxiety and reduced quality of life were observed. Preliminary research suggests that the impact of COVID-19 infection is similar.^2^

Over one million people in the UK who contracted COVID-19 report health problems more than four weeks after the onset of the acute illness. In addition, almost 700,000 people report ongoing symptoms more than 12 weeks after contracting COVID-19, a condition now referred to as ‘Long Covid’ (LC) or Post-COVID-19 Syndrome (PCS).^3^

The most common symptoms of PCS include fatigue, dyspnea, pain, anxiety and cognitive problems, but there are over 200 reported symptoms affecting ten organ systems.^4^ One study following 143 individuals seven weeks post-discharge found 53% of patients reported fatigue, 43% dyspnea and 27% joint pain.^4^ A significant number of patients report limitations with their activities of daily living (ADL), with almost 130,000 patients stating that these limitations are severe.^3^

Until recently there was no psychometrically robust patient-reported measure that focused on the impact of PCS. The COVID-19 Yorkshire Rehabilitation Scale (C19-YRS) is a 22-item patient-reported outcome measure designed to evaluate the long-term impact of COVID-19. The C19-YRS now includes clinician-completed, self-report and digital versions.^5^ Content validity of the C19-YRS has been demonstrated,^2^ and the C19-YRS is now used in the UK’s first specialist PCS community rehabilitation service,^6^ and 26 other UK National Health Service (NHS) PCS rehabilitation services. This paper describes the first stage in establishing the psychometric properties of the C19-YRS as an outcome measure for PCS.

## Methods

### Participants and Recruitment

This was a prospective observational study on patients attending a PCS Community Rehabilitation Service covering the Leeds City Region, a mixed urban and rural district in the North of England with a population of approximately 850,000 people, which includes areas of significant social deprivation. Patients were referred by their General Practitioner (GP), Community Matron or Respiratory Physiotherapy team to a PCS Community Rehabilitation Service and completed a self-report C19-YRS as part of initial triage. Data are collected in the service as part of routine clinical evaluation and ethical approval for secondary analysis of anonymized data by the University of Leeds research team was obtained (MREC 20-041).

### The C19-YRS

The C19-YRS consists of 22 items with each item scored on an 11-point numerical rating scale from 0 (none of this symptom) to 10 (extremely severe level or impact). The C19-YRS is divided into four sub-scales (range of total score for each sub-scale): symptom severity score (0 to 100), functional disability score (0 to 50), additional symptoms (0 to 60) and overall health (0 to 10). The C19-YRS was completed independently by each patient or, if the patient preferred, by a researcher or member of the administrative team via telephone. Patients’ family members or carers were permitted to help complete the responses.

On completion of clinical care for each patient, anonymized data from each completed C19-YRS was transferred to an Excel spreadsheet by clinician researchers who were not involved in the initial assessment pathway and follow-up of the patients.

### Data Analysis

Descriptive statistics are presented as arithmetic mean and SD, or median and inter-quartile range as appropriate. Absolute and relative frequencies as appropriate for demographic and categorical variables on the C19-YRS are presented. Analyses were carried out using IBM SPSS (Statistics 26, Release 26.0.0.0, 64-bit edition, IBM Corp.). Five psychometric analyses (data quality, scaling assumptions, targeting and reliability) were undertaken.

### Data Quality

Data quality concerns the extent to which a scale can be administered successfully in the target sample. The C19-YRS data were examined for percentage missing items and the percentage of the sample for whom total scores could be calculated.^7^ Imputed scores were not used for missing items.

### Scaling Assumptions

Tests of scaling assumptions examine whether it is legitimate to sum item scores to generate scale scores. In order for a set of items to be legitimately summed to form a total score, a series of criteria should be satisfied.^8-10^ We tested the C19-YRS against these criteria, which are:

1. Items should be roughly parallel, that is, measure at the same point on the scale and have similar variance, otherwise they do not contribute equally to the variance of the total score.^11^ A set of items is considered parallel when their item response option frequency distributions, and their item mean scores and standard deviations are roughly similar.^9^
2. Items should measure the same underlying construct, otherwise it is not appropriate to combine them to generate a total score.^12^ A set of items is considered to be measuring the same construct when each item’s corrected item-total correlation, which is the correlation between each item and the total score computed from the remaining items in that scale, exceeds 0.30.^11^
3. Items in the scale should contain a similar proportion of information concerning the construct being measured. This criterion is considered satisfied the corrected item-total correlations exceed 0.30.^13^

### Targeting

Targeting refers to the match between the distribution of health problems in the sample and the range of health problems measured by the scale. The better this match, the greater the potential for precise measurement. Targeting was evaluated by examining floor and ceiling effects, score distributions, and skewness statistics. Floor effects are the percentage of patients scoring zero (symptom not present) and ceiling effects are the percentage of patients scoring 10 (most severe impact of symptom) on each item. It is recommended that floor and ceiling effects should be less than 20% each on each item.^14^

### Reliability

Reliability describes the extent to which scale scores are free from random error. Scales should generate reliable estimates of the construct being measured (internal consistency). Cronbach’s alpha coefficient was used to determine this criterion.^15^ Although a range of minimum values has been suggested, it is widely accepted that Cronbach’s alpha should exceed 0.80 for group comparison studies.^10^

### Validity

Validity was evaluated by examining the extent to which Pearson’s correlations between the sub-scales of the C19-YRS were consistent with expectations.^16^ It was predicted that correlations between the symptom severity, functional disability and additional symptoms sub-scales would be moderate (between 0.3 and 0.7) and exceed the correlation with the overall health score.

## Results

### Sample Characteristics

Data for the analyses were obtained from 188 consecutive assessments of PCS patients. Patient details are given in Table 1. One patient was removed from the analyses because a significant number of answers were missing, presumed to be an oversight of the respondent.

**Table 1.** Patient demographics.

|  | **Non-hospitalised** | **Hospitalised** |
| --- | --- | --- |
| **Total no (%)** | 84% (n=157) | 15% (n=28)<br>ICU 5.4% (n = 10) |
| **Age: mean (SD)** | 47.1 (SD 13.74) | 51.9 (SD 12.83) |
| **Duration of PCS in weeks: average (SD); median (IQR)** | 24 (17); 30 (9, 38) | 18 (16); 12 (5, 33) |
| **Sex (%)** |  |  |
| Female | 66% (n=104) | 43% (n=12) |
| Male | 34% (n=54) | 57% (n=16) |
| **Ethnicity (%)** |  |  |
| White- British, Northern Irish, Irish | 80% (n=126) | 60.6% (n=18) |
| Asian/British Asian | 6% (n=10) | 18% (n=5) |
| Black – Black British, African, Black African | 2% (n=3) | 7.2% (n=2) |
| Mixed – Asian, Mixed White and Black African, Mixed White and Black Caribbean | 2.5 % (n=4) | 0 |
| Other – Undefined white, European or British | 8% (n=12) | 7.1% (n=2) |
| Arab | 1.3% (n=2) | 1% (n=3.6) |
| **Occupation (%)** |  |  |
| Healthcare workers | 30% (n=47) | 21% (n=6) |
| Non-healthcare worker | 70% (n=111) | 79% (n=22) |
| **Impact on vocation (%)** |  |  |
| Reduced hours/adjusted work | 17% (n=26) | 4% (n=1) |
| Sick leave | 19% (n=30) | 57% (n=16) |
| No effect | 64% (n=100) | 39% (n=11) |
| **Post-COVID-19 Syndrome Symptoms (%)** |  |  |
| Fatigue | 92% (n=145) | 89% (n=25) |
| Noisy breathing | 41% (n=65) | 54% (n=15) |
| Cough/throat sensitivity | 58% (n=91) | 68% (n=19) |
| Chest pain | 65 % (n=103) | 61% (n=17) |
| Muscle pain | 70% (n=111) | 68% (n=19) |
| Joint pain | 59% (n=93) | 61% (n=17) |
| Abdominal pain | 31% (n=49) | 36% (n=10) |
| Headache | 70% (n=110) | 54% (n=15) |
| Dysphagia | 23% (n=36) | 29% (n=8) |
| Faecal incontinence | 16% (n=25) | 29% (n=8) |
| Urinary incontinence | 18% (n=28) | 36% (n=10) |
| Dysexecutive syndrome | 56% (n=88) | 64% (n=18) |
| Impaired short term memory | 70% (n=111) | 75% (n=21) |
| Impaired concentration | 82% (n=129) | 82% (n=23%) |
| Depression | 70% | 75% |
| Anxiety | 92% | 90% |

Patients’ scores on the C19-YRS sub-scales are presented in Table 2. Most patients reported problems with their mental or physical health. Fatigue was the most common complaint, with 97.3% of patients reporting fatigue of varying severity, followed by the onset of pain not present before COVID-19 was contracted (94.3%). The most common new pain was muscle pain which affected 70% of patients, followed by headache (67%), chest (64%) and joint pain (59%). Approximately one third of patients also experienced new pain in their abdomen or other regions. Mental health problems were reported by 41% of patients, with 17% of these patients reporting respiratory or cardiac comorbidity. Respiratory or cardiac health issues, or both, were reported by 37% of patients. Swallowing, incontinence, skin rash and fever were bothersome for very few respondents.

**Table 2.** Patients’ scores on the C19-YRS sub-scales.

| Sub-scale (scale range) | Valid scores | Mean (SD) | Median (IQR) | Score range | Skewness |
| --- | --- | --- | --- | --- | --- |
| Symptom severity (0 – 100) | 125 | 42.7 (0.36) | 40.0 (31.0 – 54.5) | 10 – 81 | 0.232 |
| Functional disability (0 – 50) | 153 | 18.8 (10.7) | 17 (11.0 – 26.5) | 0 – 48 | 0.535 |
| Additional symptoms (0 – 60) | 155 | 18.8 (10.8) | 18.0 (10.0 – 28.0) | 0 – 48 | 0.246 |
| Overall health (0 – 10) | 183 | 4.6 (2.1) | 4.0 (3.0 – 6.0) | 0 – 10 | 0.265 |

SD = standard deviation, IQR = interquartile range; data are only presented for patients with complete sub-scale scores

### Data Quality

Missing data for items were low (range 0.5 – 19.8%). Sub-scale scores could be calculated for 67% of patients reporting symptom severity, 82% of patients reporting functional disability, 83% of patients reporting additional symptoms and 98% of patients reporting overall health. Details of scores are given in Table 2.

### Scaling Assumptions

Item response option frequency distributions were symmetric. Item means and standard deviations were similar indicating that they were roughly parallel (Table 3), although there was a greater range in symptom severity. Corrected item-total correlations exceeded 0.30 for all items except swallowing (0.24), incontinence (0.28) and skin rash (0.14) indicating that scaling assumptions were met for most items, including fever (0.33).

**Table 3.** Psychometric properties of the C19-YRS sub-scales.

|  | Symptom<br>severity | Functional<br>disability | Additional<br>symptoms | Overall health |
| --- | --- | --- | --- | --- |
| **Scaling assumptions** |  |  |  |  |
| Item means: range | 0.9 – 7.2 | 3.5 – 4.9 | 3.5 – 4.6 | 4.0 – 4.9 |
| Item SD: range | 1.9 – 3.3 | 0.3 – 1.5 | 1.1 – 1.6 | 0.8 – 1.1 |
| Item-total correlations | 0.24 – 0.62 | 0.39 – 0.67 | 0.16 – 0.62 | – |
| **Targeting** |  |  |  |  |
| Missing data (%): range | 0.5 – 19.8 | 0.5 – 15.5 | 5.9 – 12.3 | 2.1 |
| Floor effects (%): range | 5.3 - 72.7 | 16.4 – 61.0 | 15.0 – 66.8 | 2.1 |
| Ceiling effects (%): range | 0.0 – 9.6 | 0.5 – 4.8 | 0.0 – 10.2 | 1.1 |
| **Reliability** |  |  |  |  |
| Cronbach's alpha | 0.79 | 0.79 | 0.70 | – |

### Targeting

Scores spanned the range of the scale on admission and discharge and demonstrated good variability (Table 3). Results for some items demonstrated notable floor effects, especially for swallowing (72.7%), skin rash (66.8%), and fever (64.7%). There were no ceiling effects in any subscale.

### Reliability

Internal consistency of the overall C19-YRS was good (Cronbach’s alpha = 0.891). Individual sub-scales also demonstrated good reliability. Deletion of the items noted to have poor scaling assumptions and targeting improved the reliability of the symptom severity sub-scale (swallowing, incontinence removed; Cronbach’s alpha 0.79 to 0.81) and the additional symptoms sub-scale (fever, skin rash removed; Cronbach’s alpha 0.70 to 0.74).

### Validity

The symptom severity, functional disability and additional symptoms sub-scales correlated strongly with each other (Table 4), indicating that the sub-scales have a coherent internal structure. The overall health scale also correlated strongly with the other three subscales. As this is a more generic question of health status, and is an item commonly used in health-related quality of life measures, this provides some preliminary evidence of construct validity.

**Table 4.** Correlation of the C19-YRS sub-scales with the overall health scale*.

| Pearson's correlation (significance) across sub-scales |  |  |  |
| --- | --- | --- | --- |
|  | Symptom severity | Functional disability | Additional symptoms |
| Overall health | -0.322 (<0.001) | -0.352 (<0.001) | -0.208 (0.010) |
| Additional symptoms | 0.657 (<0.001) | 0.515 (<0.001) |  |
| Functional disability | 0.772 (<0.001) |  |  |

\*Overall health was reversed scored compared to item severity, so that an overall health score of '10' reflected the best possible health, in contrast to item severity where '10' reflected the worst possible severity of the symptom.

## Discussion

The C19-YRS was developed as a disease-specific patient-based measure of the impact of COVID-19 infection.^17 18^ The scale has been used successfully to gather symptom severity and functional impact and monitor progress in PCS, and is recommended by National Health Service England (NHSE)^19^ and the National Institute for Health and Care Excellence (NICE).^20^ However, it is recognized that the C19-YRS requires further iterations for development and refinement. In this first round of psychometric testing, the measurement properties of the C19-YRS were found to be good.

Many studies of rehabilitation in PCS have used generic measures of health outcome. Conceptually however, there are good arguments for making a PCS-specific scale given that many rehabilitation strategies aim to ameliorate the specific impairments associated with PCS. We examined this self-report version of the C19-YRS, initially designed for use with patients discharged from acute hospital settings, then modified to suit both hospitalized and non-hospitalized patients, to determine the stability of its psychometric properties and its potential as a measure of PCS. Our results provide evidence for that potential. In the group studied, evidence was found for data quality, scaling assumptions, targeting and reliability for most items. There is now evidence of psychometric stability across a wide sample of people with PCS. These findings from this study provide useful information and illustrate the strong potential of the C19-YRS to achieve the necessary standards for highly accurate, psychometrically robust measurement.

This study has limitations. First, it is a study from a single clinical site and includes patients with a diverse range of experiences. Whilst there is some evidence that small samples provide useful reliability and validity estimates,^21^ we recognize that our sample is relatively small at present. Nevertheless, our patient cohort is growing rapidly, and we aim to have in excess of 500 patients in our definitive psychometric analyses. Second, the scale is self-report and thus the extent to which it is applicable in patients with severe fatigue or who have impairments affecting communication remains to be determined. In this study, patients could be provided with assistance to complete the questionnaire, but is recognized that patients may answer items in questionnaires differently when the measures are self-completed compared to an interview by a member of staff, and this may lead to a bias in the reporting of the scores.^22^ Third, we have not studied test retest-reliability. However, Cronbach’s alpha is considered to be a conservative reliability estimate, and test-retest reliability often over-estimates reliability. The underpinning research for this has been discussed by Nunnally^23^ and others.^10 22 23^ Despite these limitations, we are confident that the C19-YRS will turn out to be a useful addition to current assessments of PCS in clinical studies, and could be used to complement clinician-scored measures. Furthermore, the items in the scale provide qualitative information to clinicians to assist in targeting their clinical interventions to individuals’ needs. It has advantages over other approaches, as it may be used in any setting, does not require an external rater, and is not laboratory based or require special equipment. Most importantly it measures patients’ perspectives.^24^

### Further Research

Subsequent psychometric testing will use Rasch analysis to determine whether the scale meets the fundamental axioms that define scientific measurement and permit the transformation of raw (ordinal) scores to interval level measurement.^12^ Further evaluations will examine the short- and long-term responsiveness of the scale to changes in symptom severity and the overall impact of rehabilitation on PCS. This will also determine the minimal clinically important difference (MCID) of the scale that correlates to clinical improvement or deterioration of the condition reported by patients.

## Conclusion

This is the first study to examine the psychometric properties of a PCS-specific outcome measure that captures and evaluates the symptoms experienced by patients. In this sample of patients, the C19-YRS was clinically useful and satisfied standard psychometric criteria. The C19-YRS shows good internal consistency, and scaling and targeting assumptions were satisfied. This provides preliminary evidence that the C19-YRS outcome measure of PCS has satisfactory psychometric properties.

## Data Availability

Unavailable

https://www.unavailable

## Acknowledgements

NHS England clinical guidance suggests use of the C19-YRS at first assessment, at six weeks and at six months in PCS.^19^ The National Institute for Health and Care Excellence (NICE) recommend use of the C19-YRS for comprehensive assessment of patients.^20^

This work is supported by a University of Leeds Medical Research Council (MRC) Confidence in Concept (CiC) grant for the psychometric evaluation of the C19-YRS. RJOC’s research is supported by the National Institute for Health Research (NIHR) infrastructure at Leeds and Sheffield. The views expressed are those of the authors and not necessarily those of the Medical Research Council, the NHS, NICE, the NIHR or the UK’s Department of Health and Social Care.

The C19-YRS can be freely obtained under license from the University of Leeds (https://licensing.leeds.ac.uk/product/c19-yrs-covid-19-yorkshire-rehabilitation-scale).

